# Surveillance study of acute neurological manifestations among 439 Egyptian patients with COVID-19 in Assiut and Aswan university hospitals

**DOI:** 10.1101/2020.10.28.20221879

**Authors:** Eman M Khedr, Noha Abo-Elfetoh, Enas Daef, Hebatallah M. Hassan, Mariam T Amin, Radwa K Soliman, Alaa A Attia, Amro A Zarzour, Mohamed Zain, Aliae Mohamed-Hussein, Maiada K Hashem, Sahar M Hassany, Ahmed Aly, Ahmed Shoap, Mostafa Saber

## Abstract

**Background:** COVID-19 can be accompanied by acute neurological complications of both central and peripheral nervous systems (CNS and PNS). In this study we estimate the frequency of such complications among hospital in-patients with COVID-19 in Assiut and Aswan University Hospitals.

**Material and Methods:** We screened all patients with suspected COVID-19 admitted from 1 June to 10 August 2020 to the university hospitals of Assiut and Aswan in Upper Egypt. Clinical and laboratory data, CT/MRI of chest and brain, and neurophysiology were performed for each patient if indicated.

**Results:** 439 patients had confirmed/probable COVID-19; neurological manifestations occurred in 222. Of these 117 had acute neurological disease; the remainder had non-specific neuropsychiatric symptoms such as headache, vertigo, and depression. The CNS was affected in 75 patients: 55 had stroke; the others had convulsions (5), encephalitis (6), hypoxic encephalopathy (4), cord myelopathy (2), relapse of RR-MS (2), and meningoencephalitis (1). The PNS was affected in 42 patients: the majority had anosmia and ageusia (31); the others had GBS (4), peripheral neuropathy (3), myasthenia gravis (2), or myositis (2). Fever, respiratory symptoms and headache, were the most common general symptoms. Hypertensions, Diabetes Mellitus, ischemic heart disease were the most common comorbidities in patients with CNS affection.

**Conclusion:** In COVID⍰19, both the CNS and PNS are affected. Stroke was the most common complication for CNS and anosmia and/or ageusia were common for PNS diseases. However there were 6 cases encephalitis, 2 cases of spinal cord myelopathy, 2 cases of MG and 2 cases of myositis.

## Introduction

In most patients, Covid-19 presents with a mild flu-like illness. Elderly patients with comorbidities such as hypertension, diabetes, or lung and cardiac disease, are more likely to have severe disease and deaths. Neurological complications are frequently reported in severely or critically ill patients with comorbidities. Both central and peripheral nervous systems can be affected [1, 2]. The SARS-CoV-2 virus is thought to enter the brain either via a hematogenous route [3] or via the olfactory system [4].

Most previous publications included case reports or a series of patients with stroke, encephalitis, seizures, transverse myelitis, cranial neuropathies. However in many of these, details were relatively sparse or vague (e.g. dizziness, vertigo, headache, and others), reflecting the challenge of studying such patients. A few studies showed the benefits of identifying patients with neurological complications across centres [1, 2]. A large series of 214 patients from Wuhan reported neurological symptoms in 78 patients.

There is little knowledge available concerning the acute neurological complications of COVID-19 in Egypt. This retrospective cohort study represents suspected Covid-19 patients admitted during the period from 1 June to 15 August 2020 to the university hospitals at Assiut and Aswan University in order to:

1. Estimate the proportion of acute neurological and psychiatric complications in 439 patients with COVID-19
2. The relative frequency of central and peripheral nervous system involvement.
3. The types of comorbidity associated with neurological complications.

## Material and Methods

Patients with suspected COVID-19-were admitted during the period of 1 June to 10 August 2020 at the largest two University hospitals in Upper Egypt (Assiut University Hospitals “El-Raghy hospital, Neurology, psychiatry and Neurosurgery hospital, Intensive Care Unit and Chest department” and Aswan University hospital‘” chest department, Intensive care unit and emergency unit”) as quarantine areas of isolation for those patients.

Many patients with confirmed or suspected COVID-19 had mild symptoms and were told to isolate at home. All cases admitted as inpatients had moderate to severe symptoms of COVID-19. Thus our cohort will underrepresent patients with milder symptoms, such as reduced cough, sore throat, headache, fever, fatigue, taste, smell or others. Neurologists were encouraged to report cases with neurological symptoms and signs of COVID-19 and to admit them to the Neurology, Psychiatry and Neurosurgery hospital. Computed tomography (CT) of the chest to demonstrate the ground⍰glass opacity appearance of the lungs with consolidation, laboratory investigations including, blood gases, complete blood picture, serum D-Dimer, Ferritin, C-reactive protein, renal, liver functions, coagulation profile, were obtained. For patients with neurological manifestations, CT or MRI of brain or spine, nerve conduction, EMG of upper and lower limbs were performed if needed.

**Written informed consent was obtained from each patient /or relative and Local Ethical Committee of Assiut University hospital approved the study**

### Diagnosis and Evidence of COVID-19

Reverse transcription-polymerase chain reaction (RT-PCR) is the gold standard diagnostic procedure for confirming SARS-CoV-2 virus infection. RT-PCR testing was performed on nasopharyngeal swabs using swabs with a synthetic tip, and an aluminum or plastic shaft. Swabs were placed immediately into sterile tubes containing 2-3 mL of viral transport media. The samples were stored at 2-8°C up to 72 hours.

#### RNA extraction of SARS-CoV-2

was done using the QIAamp Viral RNA Mini Kit Catalog no. 52904 supplied by QIAGEN, Germany. Sample preparation using QIAcube instruments follows the same steps as the manual procedure (i.e., lyse, bind, wash and elute).

#### Pathogen detection of SARS-CoV-2 RNA

used the TaqMan™ 2019-nCoV Control Kit v1 (Cat. No. A47532) supplied by QIAGEN, Germany on the Applied Biosystem 7500 Fast RT PCR System, USA. It amplified and detected three viral genomic regions, reducing the risk of false negatives including the N protein (nucleocapsid gene), S protein (Spike gene), and open reading frame-1ab (ORF1ab) genes. Applied Biosystems™ TaqMan™ 2019-nCoV Control Kit v1 (Cat. No. A47533) is a synthetic positive control that contains target sequences for each of the assays included in the TaqMan™ 2019-nCoV Assay Kit v1 (Cat. No. A47532).

### Evidence of SARS-CoV-2 infection was defined as

1. definite COVID-19 if patients came with clinical symptoms of infection and had positive PCR of respiratory samples (eg, nasal or throat swab).
2. probable COVID-19 if clinical symptoms of infection and chest CT were consistent with COVID-19 and one or 2 laboratory investigations were positive (lymphopenia, high serum ferretin and D-Dimer Level) but PCR was negative or not done.

### Diagnosis of Neurological disorders

When patients met more than one specific clinical case definition (eg, seizures and encephalitis or stroke), the underlying causal diagnosis was considered primary and complications of that diagnosis considered as secondary features (eg, encephalitis or cerebrovascular stroke would be considered primary and seizures secondary).

### Definition of major neurological diseases

#### Cerebrovascular disease

Stroke was defined according to WHO criteria as a syndrome of rapidly developing clinical signs of focal (or global) disturbance of cerebral function, with symptoms lasting 24 h or longer or leading to death, with no apparent cause other than of vascular origin [5], and documented by CT brain as ischemic or hemorrhagic stroke.

#### Subarachnoid hemorrhage

Bleeding into the subarachnoid space as documented by CT brain scan. The classic presentation can include: sudden onset of severe headache (the classic feature), accompanying nausea or vomiting, symptoms of meningeal irritation, photophobia and visual changes, focal neurologic deficits and sudden loss of consciousness at the ictus.

#### Cerebral venous sinus thrombosis (CVST)

The classic presentation can include the following: headache, blurred vision, fainting or loss of consciousness, loss of control over movement in part of the body, seizures and coma.

#### Encephalitis

Symptoms include high fever, headache, sensitivity to light, stiff neck and back, vomiting, altered mental state, confusion and, in severe cases, seizures (clinical or electroencephalographic evidence), paralysis and coma with exclusion of iatrogenic factors, such as sedatives and antipsychotics. Encephalitis documented by the presence of brain edema and signs of inflammations in MRI.

#### Hypoxic Encephalopathy

Altered sensorium ranging from confusion, delirium, stupor to coma due to prolonged hypoxia with exclusion of iatrogenic factors, such as sedatives and antipsychotics.

#### Meningitis

Symptoms such as headache, fever and a stiff neck and manifestation of increase intracranial tension.

#### Transverse myelitis

Transverse Myelitis ™ is a disorder caused by inflammation of the spinal cord. It is characterized by symptoms and signs of neurologic dysfunction in motor and sensory tracts on both sides of the spinal cord. The involvement of motor and sensory control pathways frequently produce altered sensation, weakness and sometimes urinary or bowel dysfunction.

#### Guillain–Barre’ syndrome (GBS)

GBS is defined as an acute illness characterized by clinical symptoms beginning with progressive ascending weakness (usually beginning peripherally in the limbs), impairment of position and vibration sense, reduced or absent tendon reflexes and confirmed by prolonged distal latencies and F wave and reduction of motor conduction velocities.

### Statistical analysis

The collected data was revised, coded, tabulated and introduced to a PC using Statistical package for Social Science (SPSS 25). Data was presented and suitable analysis was done according to the type of data obtained for each parameter.

Number and percent or Means ± standard deviation (SD) were used to represent data. The level of significance was set at *P* < 0.05. Patients with neurological manifestations were classified into 3 groups: Patients with acute CNS disease (number =75), patients with acute PNS disease (number = 42 cases), and patients with non-specific neuropsychiatric manifestations. (number=105).

## Results

Out of 439 patients with confirmed /probable COVID-19, 222 (50.6%) had neurological manifestations. 117 (52.7%) of these had acute neurological disease: 75 (64.1%) with central neurological disorders and 42 (35.9%) with acute peripheral neurological disorders; the remaining 105 patients had non-specific symptoms such as anxiety, dizziness, headache, depression and anxiety (Fig 1). The most common CNS disease was cerebrovascular disease (55 patients (73.3%), most of them were ischemic stroke (42 cases: 9.6% of 439) while 13 patients (3% of 439) had hemorrhagic CVD. There was one case each (0.22%) of subarachnoid hemorrhage, subdural hematoma with intracerebral hemorrhage out of patients with cerebral hemorrahge.

**Figure 1.**
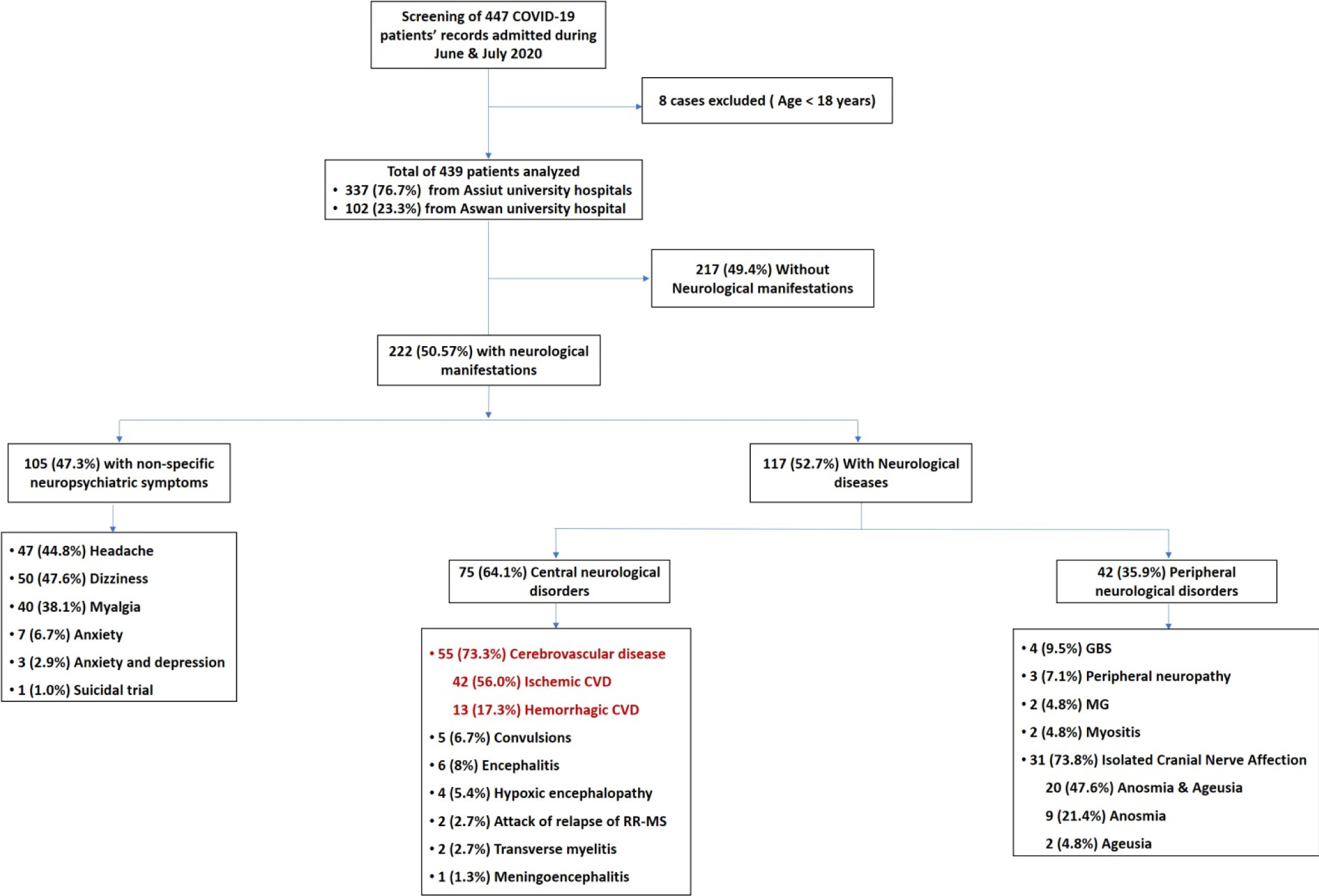
Flow chart of studied patients

Encephalitis was recorded in 6 patients (8%), followed by convulsions (5 patients (6.7%)), hypoxic encephalopathy (4 patients (5.4%)), transverse myelitis (2 patients (2.7%)), relapse of multiple sclerosis (RR-MS (2 patients (2.7%)) and meningoencephalitis (1 patient (1.3%)). Of the 41 cases with PNS disease, 31 (73.8%) had isolated cranial nerve affection (anosmia and/or ageusia), 4 (9.5%) had GBS, 3 (7.1%) had peripheral neuropathy, 2 (4.8%) had myasthenia gravis, and 2 (4.8%) had myositis (Figure 1 Flow chart)

Table 1 details the demographics and symptomatology of the patients. Patients with acute CNS disorders were significantly older than the other groups. There was significant difference between the percent of definite/probable COVID-19 between patients with CNS disorders versus those with PNS disorders. The most common presenting symptoms were fever, respiratory symptoms and signs and headache followed by fatigue and malaise, GIT symptoms, dizziness, and/or vertigo; the least frequent symptom was sore throat. The percentage symptoms was significantly higher in patients with PNS because most of the patients with acute CNS disease had disturbed consciousness making it impossible to asses them in detail (most of them had cerebrovascular stroke).

**Table 1:** Patient details and results of chest CT

| Variables | Total number of patients with definite neurological disease N=117 | Patients with acute CNS disease N=75 | Patients with acute PNS disease N=42 | P value |
| --- | --- | --- | --- | --- |
| Age Mean + SD | 55.1±17.2 | 59.5±17.5 | 48.3±14.3 | 0.001 |
| Sex M/F Ratio | 61/56<br>52.1%/47.9% | 41/34<br>54.7%/45.3% | 20/22<br>47.6%/52.4 % | 0.600 |
| Definite COVID-19<br>Probable COVID-19 | 55(47.0%)<br>62(53.0%) | 23(30.7%)<br>52(69.3%) | 32(76.2%)<br>10(33.8%) | 0.0001 |
| Presenting of general symptoms |  |  |  |  |
| Fever | 101(86.3%) | 70(93.3%) | 31(73.8%) | 0.003 |
| Sore throat | 7(6.0%) | 0 (0%) | 7 (16.7%) | 0.0001 |
| Fatigue and malaise | 30(25.6%) | 14 (18.7%) | 16(38.1%) | 0.003 |
| Headache | 36(30.8%) | 21(28%) | 15(35.7%) | 0.380 |
| Dizziness, and/or vertigo | 15(12.8%) | 4(5.3%) | 11(26.2%) | 0.001 |
| GIT symptoms and signs | 29(24.8%) | 17(22.7%) | 12(28.6%) | 0.321 |
| Respiratory symptoms and signs | 100(85.5%) | 60(80.0%) | 40(95.2%) | 0.781 |
| CT chest: Positive bilateral/unilateral GGO | 98(83.8%) | 69(87.3%) | 29(69%) | 0.043 |

PCR; Polymerase chain reaction, GIT ; gastrointestinal symptoms, GGO; ground glass opacities with consolidation, CT chest; Computed tomography chest

Table 2 indicates that 79 (67.5%) cases out of 117 had one or more comorbidity risk factors; these were significantly more common in patients with CNS disorders than PNS disorders, particularly renal impairment and previous neurological disease (particularly CVS). In general the commonest comorbidities were hypertension followed by diabetes and ischemic heart disease and neurological disease.

**Table 2:** Comorbidities among studied groups

|  | Total number<br>117 cases | Patients with<br>acute CNS<br>disease<br>N=75 | Patients with<br>acute PNS<br>disease<br>N=42 | P value |
| --- | --- | --- | --- | --- |
| **Comorbidities** | 79(67.5%) | 57 (76%) | 22 (52.4%) | 0.008 |
| Hypertension | 53 (45.3%) | 38(50.7%) | 15(36.7%) | 0.119 |
| Ischemic heart | 20(17.1%) | 14(18.7%) | 6(14.3)) | 0.546 |
| Diabetes Mellitus | 41(35%) | 25(33.3%) | 16(38.1%) | 0.605 |
| Impaired liver function | 6(5.1%) | 6(8%) | 0 | 0.218 |
| Impaired renal function | 10(8.4%) | 9(12%) | 1(2.4%) | 0.074 |
| Chest disease | 4(3.4%) | 2(2.7%) | 2(4.8%) | 0.550 |
| Neurological disease (Types) * | 14(12.0%) | 11(14.7%) | 3(7.1%) | 0.229 |

\*ischemic stroke, hemorrhagic stroke, epilepsy, PD, PD and AD, AD, Brain tumor, MG, CKD and PD.
PCR; Polymerase chain reaction, GIT ; gastrointestinal symptoms, GGO; ground glass opacities with consolidation, CT chest; Computed tomography chest

The mean serum level of D dimer was 2.737± 6.63 (range 0.19-35.2). A significantly higher level was seen (8.77±11.2: range 0.46-35.2) in patients with acute CNS disease than with acute PNS disease (0.79±0.92: range 0.19-3.6) (p=0.0001). Mean serum ferritin was 544.7±723.4 (range 10-3859). It was non-significantly higher in patients with acute CNS disease (787.2±1204.3: range 143-3959) than in patients with PNS disease (463.9±480.6: range 10-1819.7) (p=0.251). The mean percent lymphocytes was 18.6±12.5 (range 2.5-59.3). Patients with acute CNS disease had a significantly lower level (16±12.3: range 2.5-59.3) than in PNS disease (23.1±11.8: range 7.98-49.8) (p=0.003).

## Discussion

Available pieces of evidence suggest that the SARS-CoV-2 virus can traverse the blood–brain barrier and enter the brain, possibly via a hematogenous route. It may also enter trans-neuronally via the olfactory system, across the cribriform plate [4]. Angiotensin-converting enzyme 2 receptors that are present on endothelial cells of cerebral vasculature act as cell entry points for virus [3].

### Cerebrovascular disease

Many cerebrovascular events were identified in our study (55 cases (73.3%) out of 75 patients with CNS symptoms; or 12.5% out of the whole sample of 439 COVID patients). This is consistent with previous cohorts and case reports of acute COVID-19 complications [6, 2, 7]. Most of them were ischemic stroke (42 cases: 9.6% of 439) while 13 patients (3% of 439) had hemorrhagic CVD, out of them one case had subarachnoid hemorrhage, and another had subdural hematoma with intra-cerebral hemorrhage. Li and co-workers[8], in a retrospective study, noted that out of 221 patients, 13 (5.9%) developed cerebrovascular disease after infection. Of these patients, 11 (84.6%) were diagnosed with ischemic stroke, 1 (7.7%) with cerebral venous sinus thrombosis, and 1 (7.7%) with cerebral hemorrhage. In a case series of 12 patients, [9] Reddy S.T. et al found that 10 patients had an ischemic stroke, of which 1 suffered hemorrhagic transformation and two had intracerebral hemorrhage. [6] Beyrouti and co-workers, in a report of six severely affected patients with large cerebral infarcts, noted elevated D-dimer levels (≥1000 µg/L), indicating a coagulopathy. Few other studies have reported cerebrovascular complications in COVID-19 [10, 2]. Other small case series have described patients with COVID-19 and concurrent stroke [11, 7].

The pathophysiological mechanisms that underlie cerebrovascular ischemic events in COVID-19 could potentially be related to vasculopathy, with a report of SARS-CoV-2 endothelitis-related cerebrovascular events [12]. In addition, there is an increase in conventional stroke risk during sepsis [13, 14]. Comorbidities, such as diabetes and hypertension enhance expression of the angiotensin-converting enzyme 2 receptor in the brain and neurotropism of the SARS-CoV-2 virus[15]. In COVID-19, alterations in blood pressure control are another proposed mechanism that has been suggested to explain the increased risk of cerebral vascular complications. Ordinarily, angiotensin-converting enzyme 2 signaling lowers blood pressure[16]. Competitive blockage of angiotensin-converting enzyme 2 by the SARS-CoV-2 virus down-regulates angiotensin-converting enzyme 2 expression leading to uncontrolled blood pressure and the enhanced possibility of cerebrovascular accidents.

### Cord Myelopathy

Spinal cord involvement is rare in COVID⍰19. In the present study we reported two cases with spinal cord involvement out of 439 cases (0.5%) with COVID-19. The first patient came with acute onset of flaccid paraplegia and a sensory level at T4 associated with retention of urine, 10 days after flu like symptoms. The MRI revealed a picture of transverse myelitis. The second patient presented with acute quadriplegia with preservation of deep sensation, and a history of fever, headache and insomnia 3 days previously. The MRI showed cervicodorsal myelopathy of the anterior 2/3 of the cord extending from C4-T4 most probably caused by secondary occlusion of the anterior spinal artery following acute COVID-19 pneumonia. Only 4 cases of transverse myelitis have been reported previously [17-20]. It is probably due to exaggerated inflammatory changes following a cytokine storm brought on by Covid-19 [20].

### Hypoxic Encephalopathy

There were 4 confirmed (nasopharyngeal swab) Covid-19 cases (0.25% of the total or 5.4% of those with CNS symptoms) who presented with altered sensorium ranging from confusion, delirium, stupor to coma due to prolonged hypoxia and low oxygen saturation. They came to the hospital complaining of headache, altered mental status, fever, and cough. Upon examination, the patients were found to be encephalopathic, not responding to any commands. Non-contrast, brain CT scan showed no acute abnormalities whilst electroencephalography (EEG) showed bilateral mild diffuse slowing of background activity.

### Encephalitis

The SARS⍰CoV⍰2 virus has the potential to enter the brain by travelling from the nasal mucosa to the olfactory bulb and spreading to the piriform cortex. Xiang and co⍰workers [21] in Beijing, China, claimed to isolate the first SARS⍰CoV⍰2 virus in cerebrospinal fluid (CSF). So far there have been seven additional reported cases of SARS⍰CoV⍰2 associated encephalitis, encephalopathy, or meningitis. In the present study, six cases had features consistent with encephalitis. The SARS⍰CoV⍰2 virus was identified in a throat swab in four cases while the other 2 cases were probable Covid-19 with positive chest CT that demonstrated the ground⍰glass appearance of the lungs and leukocytosis and lymphopenia. Neuroimaging was normal. Unfortunately we were unable to examine the CSF.

### Seizures

Seizures are not a common manifestation in COVID-19. In the present study there were 19 cases out of 439 (4.3%). In 3 of them (0.68%) the seizures were novel; another 2 patients had a known history of controlled epilepsy and had experienced exacerbation of the seizures with COVID-19 (0.46%). In the other 14 patients (3.19%) the seizures were associated with other primary pathology (5 patients had recent ischemic stroke, 2 patients had recent hemorrhagic stroke, 6 patients had encephalitis and 1 patient had an old brain tumor). All our cases were diagnosed on clinical and electroencephalographic evidence. In contrast to our results, a multicentric Chinese retrospective study noted that among 304 COVID-19 patients (108 with severe disease) none had acute symptomatic seizures or status epilepticus despite the presence of severe metabolic alterations. Mao and co-workers [2] noted that among 214 patients admitted to intensive care units there was only a single case of seizure. Vollono and colleagues [22] published a report of nonconvulsive status epilepticus triggered by Covid-19, in an elderly patient. The higher frequency of seizures in Covid-19 patients in our study may be related to the omission of mild cases and specialty admission of cases with neurological manifestations to the Neurology, Psychiatry and Neurosurgery department, Assiut University.

### Guillain-Barré syndrome (GBS)

Post-infectious autoimmune reactions can affect neuronal cells. The SARS-CoV-2 virus epitopes bear a structural resemblance to several human proteins. Molecular mimicry between virus epitope and myelin basic protein results in autoimmune post-infectious demyelinating syndromes[23].

In the present study, GBS is a frequently encountered neurological complication of COVID-19. Four cases presented with classical clinical manifestations of acute symmetric flaccid weakness with absent reflexes and numbness of four limbs, 2 to 3 weeks after Covid-19. Nerve conduction, F waves, H reflex and electromyography confirmed the presence of severe demyelinating polyradiculoneuropathy. Zhao and co-workers [24] described the first patient of Guillain-Barré syndrome in a patient with COVID-19.

### Myositis and covid-19

In the present study, 2 (4.8%) cases had myositis. The firstpatient came to the hospital with a 3-4 day history of fever, lower respiratory tract symptoms (cough, sputum, and dyspnea). Then he developed respiratory distress, bulbar symptoms, neck and proximal muscle weakness and pain of all four limbs with no sensory impairment. PCR was positive, for Covid-19 with hypoxemia, anemia, leukocytosis, lymphopenia, with high creatine kinase (CPK), ferritin levels and D-dimer. A chest CT showed bilateral Ground glass opacity. The patient needed non-invasive mechanical ventilation. Electromyography (EMG) confirmed the clinical diagnosis of myositis. The 2^nd^ patient had a 7 day history of fever and cough. She developed severe muscle pain and weakness of four limbs, and EMG confirmed the clinical diagnosis of myositis. Her laboratory data showed decreased PCO2, anemia, leukocytosis, neutropenia and lymphopenia with high CPK and ferritin. CT chest showed bilateral ground glass opacity. PCR was not performed.

Zhang et al [25] reported a patient with COVID-19 associated inflammatory myopathy, presenting with facial, bulbar and proximal limb weakness. Beydon and co-workers [26] recently described myositis in a critically ill patient of COVID-19. The patient presented with acute myalgia, difficulty in waking, and proximal weakness. CPK (25384 IU/L) were markedly elevated. Four days later, the patient became febrile and tested positive for the SARS-CO-V-2 virus.

### Myasthenia Gravis (MG) and covid-19

In the present study two patients with Covid-19 developed MG (4.8%) and required intubation for respiratory failure. One patient with previously stable MG had exacerbation of myasthenia with bilateral facial, bulbar, masticatory symptoms and proximal weakness of four limbs. The second previously well patient developed MG with ptosis, bulbar manifestations and proximal muscle weakness. The diagnosis was confirmed by a positive decrement test in neurophysiology study and elevated serum acetyl choline receptor antibody in both cases. Both patients were treated with intravenous immunoglobulin and improved. To our knowledge only one study of Anand et al [27] reported five hospitalized patients with autoimmune MG (four with acetylcholine receptor antibodies, one with muscle specific tyrosine kinase antibodies)

### Smell and taste

In the present study, 42 cases had PNS disease. The most frequent (31 cases, 73.8%) symptoms were a complete or partial loss of smell (anosmia) and taste (ageusia) consistent with recent reports of Moein et al [28]. In a French study, Lechien and co⍰workers [29] reported that out of 417 mild⍰to⍰moderate COVID⍰19 patients, 86% and 88% of patients, respectively, reported anosmia and ageusia. The SARS⍰CoV⍰2 virus utilizes angiotensin⍰converting enzyme 2 receptors, present in the olfactory epithelium, to enter into the neuronal cells from where it spreads to the olfactory bulb via the olfactory nerve [30].

### General symptoms

In the present study fever occurred in 101/117(86.3%) patients with neurological complications with a higher frequency among patients with acute CNS vs PNS disorders (88.6% vs 73.8%). This is consistent with other meta-analyses that reported fever as a common general manifestation in COVID-19 patients with neurological manifestations, ranging from 80.4% to 88.7% [31-34].

Respiratory symptoms including cough with or without expectoration and dyspnea were also recorded in the majority of cases (100/117patients) but were more frequent among patients with acute PNS than acute CNS diseases. Thirty-nine patients reported GIT symptoms. Respiratory symptoms were more common in the present study than in other studies [31-34] ranging from 57-63% for cough and 45-58% for dyspnea.

Fatigue and malaise were observed in 30/117 (25.6%) patients and headache in 36/117 (30.8%) patients; both were more common in PNS diseases than in CNS diseases. Our recorded rates were lower than those in some previous meta-analyses (38-46% for fatigue) [32, 34]. In 1,420 mild-to-moderate Covid-19 patients, headache (70%) was the most prevalent symptom [35]. On other hand our recorded rate of headache was higher than recorded rate (8-15%) in other studies [32-34]. Increased mental stress, excessive anxiety, and changes in lifestyle are possible reasons for early headaches. Belvis R [36] in a recent communication opined that COVID-19-associated acute headache can be due to systemic viral infection, primary cough, headache, and tension-type headache. Early headaches respond well to acetaminophen. Headaches appearing between the 7th and the 10^th^ days of illness may be related to cytokine storm [36]. Vertigo and dizzeness were observed in (15/117 patients), while sore throat was the least frequent symptom (7 /117 patients).

## Conclusions

Covid-19-associated acute neurological diseases are common in our study. Acute CNS diseases were more common than PNS diseases. Cerebrovascular stroke was the most common CNS diseases and loss of smell and taste were the most common PNS diseases. Comorbidities were common among patients with CNS diseases. These data begin to characterize the spectrum of neurological complications of COVID-19 in Egypt.

## Data Availability

all the data of the manuscript available on request

